# Cerebral white matter integrity in Type-2 Diabetes Mellitus and integrated role of Interleukin-6: Findings in multiethnic middle-aged and older US-adults

**DOI:** 10.64898/2026.11.02.26367084

**Authors:** Asma Hallab, The Health and Aging Brain Study (HABS-HD) Study Team

## Abstract

**Introduction:** Peripheral neural injuries, known as polyneuropathy, are largely described in patients with Type-2 Diabetes Mellitus (T_2_DM). However, less is known about the T_2_DM-associated cerebral White Matter (WM) integrity. Previous studies highlighted mediating effects of inflammation in the T_2_DM-brain association. It is, however, still unclear whether Interleukin-6 (IL-6) mediates specifically T_2_DM–cerebral WM integrity.

**Methods:** The multiethnic ≥50-year-old study population is a subset of the Health and Aging Brain Study: Health Disparities (HABS-HD) study. Diffusion-weighted scans were analyzed, and diffusion parameters were averaged in major WM tracts. Effect sizes (Cohen’s *d*) were calculated after adjusting for relevant confounders. Mediation analysis was performed using 1000-fold non-parametric bootstrapping of 95% confidence intervals (CI). The results were adjusted for multiple testing using the False Discovery Rate method (FDR).

**Results:** Several WM tracts showed significant impairment in T_2_DM participants after adjusting for confounders and multiple testing (posterior thalamic radiation (adj. Cohen’s *d*=0.14[0.03-024]), anterior corona radiata (adj. Cohen’s *d*=0.15[0.04-0.26]), cingulum (adj. Cohen’s *d*=0.14[0.03-0.25]), inferior fronto-occipital fasciculus (adj. Cohen’s *d*=0.16[0.06-0.27]), genu of the corpus callosum (adj. Cohen’s *d*=0.13[0.03-0.24]), and body of the corpus callosum (adj. Cohen’s *d*=0.28[0.17-0.38])). Within two years, the effect sizes showed an increasing trend. Even in cognitively unimpaired participants, T_2_DM was associated with locally reduced WM integrity. IL-6 statistically mediated up to 15% of these major associations.

**Conclusions:** The study demonstrated T_2_DM-associated impairment of intracerebral WM integrity. In T_2_DM, microstructural WM changes might start before clinically objectified cognitive impairment. The role of systemic inflammation needs to be assessed in causal frameworks.

## 1. Introduction

Type-2 Diabetes Mellitus (T_2_DM) has shown a pandemic trend in the last few decades, mainly owing to the changes in lifestyles, eating habits, and increased obesity risks. (1–3) Not only do insulin resistance and metabolic disorders mark the seriousness of the disease, but also the chronic and irreversible damage caused to several organs. These associate T_2_DM with higher morbidity and mortality rates, particularly in the early-onset subgroup. (4–6) Data on different US communities demonstrated higher risk profiles in Hispanic and Black communities than those recorded in non-Hispanic White ones. (7) Mexican and African Americans in particular, have an earlier age of onset of T_2_DM, higher psychological burdens, and higher inflammatory profiles. (8) These risk factors might increase their overall vulnerability to health complications and amplify their risks of earlier disease-related mortality. (8–10) On similar scales, epidemiological studies reported alarming records on the worldwide increase of dementia incidence. It is estimated that 152 million of the world population will develop some form of dementia by 2050. (11) In addition to the primary cause associated with aging and the growth of the global population, these high rates are also attributed to an increasing incidence of glucose metabolism disorders. (12, 13)

Our previous study showed a significant association of T_2_DM with amyloid, Tau, and neurodegeneration (ATN) biomarkers at preclinical stages of diabetes-related cognitive impairment. (8) In cognitive testing, the association involved mainly executive function, flexibility, and decision-making as measured by the Trail-Making-Test-B (TMT-B). (8) The domain-specificity theory of T_2_DM-associated adversities is supported by another study, based on the same cohort and restricted to cognitively unimpaired participants. This showed a significant correlation between higher glycated hemoglobin A1 (HbA_1c_) levels and regionally thinner cortex, particularly in temporal, parietal, occipital areas, and some frontal regions (14) in addition to hippocampus neurodegeneration. (8)

While insulin resistance’s role remained a valid causality-driven hypothesis in several previous studies, low-grade inflammation also appears to be a significant actor in these associations. Interleukin-6 (IL-6) mediated around 4 to 12 % of the association between long-lasting diabetes, and amyloid ß, total tau, neurofilament light chain (NfL) levels, and hippocampal volume. (8) Furthermore, in patients with T_2_DM, the lowest insulin sensitivity was significantly associated with the highest levels of pro-inflammatory biomarkers, mainly IL-6. (15) This theory was further supported by pre-clinical studies, where IL-6 was a significant mediator between peripheral metabolic alterations and Alzheimer’s disease (AD) pathology. (16)

In our previous study, Neurofilament Light Chain (NfL) showed a powerful association with T_2_DM and IL-6, unveiling an underlying active state of neural injury. (8) However, high plasma NfL levels are derivatives of both central and peripheral axonal injuries, including those described in diabetic polyneuropathy. (17, 18)

The T_2_DM-associated intracerebral adversities have long been classified as microvascular complications. (19, 20) It is, however, unclear whether inflammation might play a mediating role in the effect of T_2_DM on cerebral WM integrity, comparable with the intracerebral neural injuries described in our previous study. (8) It is also well known that inflammation-associated neuropathy is rather of a demyelinating nature, and the combination of T_2_DM and IL-6 effects on intracerebral WM needs to be better understood.

The main hypothesis of the study is that participants with T_2_DM have impaired WM integrity compared to those without T_2_DM. The second hypothesis is that IL-6 plays a mediating role in this association.

The aim of the current study was to explore the association between diffusion tensor imaging (DTI)-measured WM integrity biomarkers and T_2_DM in community-dwelling middle-aged and older US adults with equally represented ethnic groups and to understand the statistical mediating role of IL-6 in this constellation.

## 2. Methods

The Strengthening the Reporting of Observational Studies in Epidemiology (STROBE) guidelines were followed for the elaboration of this manuscript. (21)

### 2.1. Study population

The study population is a subset of the Health and Aging Brain Study: Health Disparities (HABS-HD), a monocenter study performed in the Institute for Translational Research at the University of North Texas Health Science Center (UNTHSC) in Fort Worth, Texas - USA, where participants from North Texas were recruited. The study’s first aim was to assess differences in health outcomes between White and Mexican Americans (HABLE, initiated in 2017). (22) From 2020, further 1,000 African American participants were added to the source cohort. (23) The target population was multiethnic community-dwelling middle-aged and older adults, and participants were recruited through media and in community events. Participants were eligible if they were at least 50 years old, had never been diagnosed with any type of dementia (except Alzheimer’s), and had no history of substance use disorders. Participants with Type-1 Diabetes Mellitus (T_1_DM), history of severe Traumatic Brain Injury (TBI), recent cancer (current or during the last 12 months), severe mental illness (except depression and anxiety), or advanced stages of chronic physical conditions that might impact their cognition (severe chronic kidney disease or heart insufficiency…) were not eligible. Having an acute active infectious status was also an exclusion criterion. Participants were interviewed by certified clinicians, including mental health professionals and neuropsychologists. They underwent clinical, neuropsychological, biological, neuroimaging, and genetic assessments. (23)

The study was based on the 6^th^ data release of HABS-HD.

All participants gave written informed consent. HABS-HD was approved 12/2/2025 by the North Texas Regional Institutional Review Board, Reference #2016-128. All procedures contributing to this work comply with the ethical standards of the relevant national and institutional committees on human experimentation and with the Declaration of Helsinki 1975, as revised in 2013. The current research is based on a secondary analysis of anonymized data and was performed in compliance with the data use agreement.

### 2.2. Type-2 Diabetes Mellitus

For the current analysis, T_2_DM was diagnosed if the participants disclosed a history of T_2_DM under current medical therapy (Insulin, oral medication, special diet) or when the glycated hemoglobin A1c (HbA1c) levels were equal to or higher than 6.5%. Furthermore, fasting blood glucose levels (mg/dL) were reported.

### 2.3. Neuroimaging

Participants underwent 3 Tesla diffusion magnetic resonance imaging (dMRI) scans; 1,620 (76%) underwent with Siemens Magnetom Skyra and 524 (24%) Siemens Magnetom Vida 1, following the first DTI protocol– HABLE/HABS-HD, including b=0 and b=1000 s/m^2^ shells. Neuroimaging data were analyzed at Shi’s Laboratory following the lab’s pipeline. dMRI scans underwent motion and Eddy current correction using the FSL tool (v 6.0.7, FMRIB, Oxford, UK). (24) Following, dMRI scans underwent a skull stripping and were resampled to an isotropic resolution of 2 mm^3^. dMRI parameters, mainly Fractional anisotropy (FA), were computed using MRtrix3. (25) For each participant, a mean value of FA was calculated across 32 white matter tracts defined by the JHU-MNI Type-II Atlas. (26)

### 2.4. Interleukin-6

Single Molecule Array (SIMOA) technology, developed by Quanterix™, is a highly sensitive method that was used to measure IL-6 in fasting blood plasma. IL-6 levels were reported in pg/mL. (22)

### 2.5. Neuropsychological tests

Included neuropsychological test scores were: Geriatric Depression Scale (GDS), (27) Mini- Mental-State Examination (MMSE), (28) and Trail-Making-Test-B (TMT-B). (29)

### 2.6. Inclusion criteria

Only participants with complete information on their T_2_DM status, IL-6 levels, and processed dMRI scans were included in the analysis. As previously mentioned, participants with T_1_DM or clinically relevant acute infection were not eligible.

### 2.7. Confounders

The models were adjusted for: age (years), sex (male vs. female), self-disclosed race/ethnicity (“White” vs. “Hispanic” vs. “Black”), education (years), Apolipoprotein E (APOE) ε4 positivity (none vs. at least one allele), cognitive status (“Healthy Control” (HC), “Mild Cognitive Impairment” (MCI), and dementia), Body Mass Index (BMI = Weight (Kg) / Height (m)^2^), cardiovascular diseases (CVD) (binary), hypertension (binary), dyslipidemia (binary), current smoking (binary), alcohol consumption (binary), and type of MRI scanner used (Skyra vs. Vida 1). Additionally, models were adjusted for the use of anti-inflammatory medication (AIM).

As in previous publications, (30, 31) the AIM was classified after filtering case-by-case the medication list and extracting relevant information:

- All non-steroidal anti-inflammatory drugs (NSAIDs).
- All systemic corticoids (unless details on the administration route are missing).
- All disease-modifying anti-rheumatic drugs (DMARDs) (for rheumatological and systemic disorders, and including a few cases with post-transplantation medication).

### 2.8. Statistical analysis

Variables were visualized and analyzed for completeness and distribution using plotting and the Shapiro-Wilk test. Less than one percent of BMI (n=13) and APOE ε4 (n=9) values were missing at random and were imputed by the median and null, respectively. IL-6 levels were right-skewed and were log-scaled for the analysis. Differences in participants with and without T_2_DM were assessed using the Wilcoxon rank sum test, Pearson’s Chi-squared test, and Fisher’s exact test. The same methods were used to compare participants at the 2-year follow-up.

An exploratory Spearman’s rank correlation was applied to assess the correlation strength of the immunometabolic (particularly IL-6) and neurocognitive biomarkers with WM FA in different tracts, bilaterally. Variables were visualized in a correlation matrix. Seeing the high correlation coefficients (r=1) between the right and left structures (Figure 1), and an expected lack of laterality effect in the association between T_2_DM and WM tracts in the primary hypothesis, a mean FA value was calculated by averaging the FA on the right and the FA on the left structure. This method is largely described in the literature. (32, 33)

**Figure 1:**
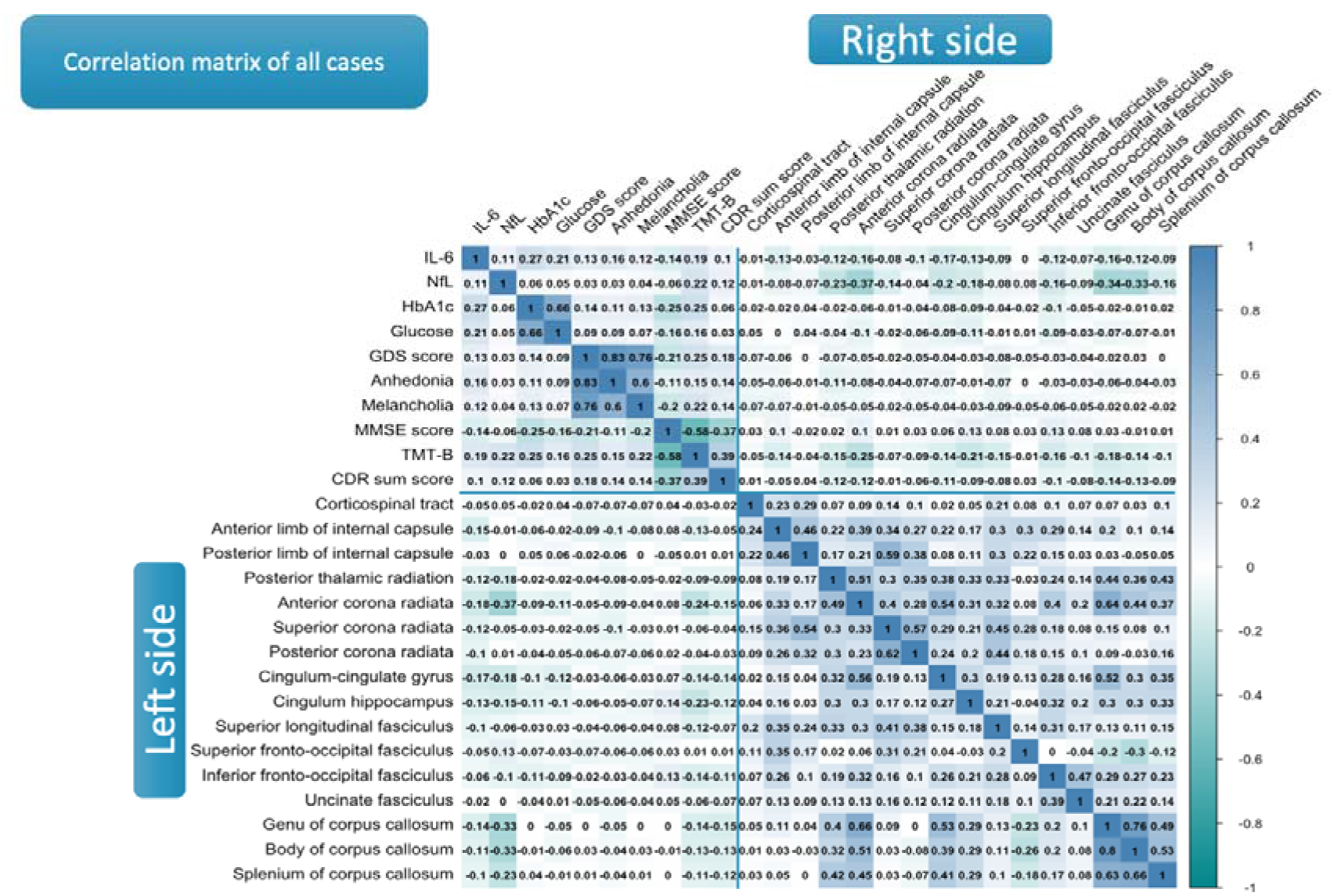
Correlation matrix of immunometabolic biomarkers and fractional anisotropy in different white matter tracts – A bilateral approach: Total study population

Then, a detailed matrix showing only significant correlation coefficients was created in both subpopulations to quantitatively estimate an indirect biomarker of WM connectivity in each group.

Linearity assumptions were tested for all models where the FA of WM tracts was the outcome, T_2_DM was the predictor, and all covariates were included.

Model-based adjusted means were computed using the Estimated Marginal Means method. The difference in adjusted mean FAs between participants without and with T_2_DM was computed by subtraction (non-T_2_DM minus T_2_DM). A model-based adjusted effect size (Cohen’s *d*) was then derived from the results. To ensure the correctness and comparability of the findings, a standardized regression coefficient (*ß*) was also reported. The confounders included in Cohen’s *d* and standardized regression coefficient equations are detailed in 2.6. Significant results were adjusted for multiple testing using the false discovery rate (FDR) method in order to reduce Type I error risk. All *p*-values resulting from different tract comparisons (Adjusted *p*-value*) were included in the same FDR equation (n=16; Adjusted *p*_FDR_-value). The significant results defined the “T_2_DM-specific WM tracts” for this study. The first sensitivity analysis was based on assessing differences between non-T_2_DM and T_2_DM regarding WM FAs across all tracts at 2-year follow-up using the same earlier described method. Then a second sensitivity analysis was performed on cognitively unimpaired participants at baseline.

The association between T_2_DM and WM integrity in “T_2_DM-specific WM tracts” was assessed using a multivariable linear regression analysis, including the pre-described confounders (2.6.). A confounder-adjusted non-parametric mediation analysis with 1000-fold bootstrapping of a 95% confidence interval (CI) was performed on significant associations in order to assess the mediating role of IL-6. Results were calculated as the proportion of mediated effect on the total effect and presented as percentages (%) with 95% CI.

RStudio version 2025.05.1+513 (Posit Software^©^, PBC, USA) was used for the statistical analysis.

## 3. Results

### 3.1. Study population

From a total of 3,838 participants, 1,649 were missing IL-6 levels and 220 were missing neuroimaging variables at the analysis timepoint. A total of 2,144 participants fulfilled the inclusion criteria.

A comparable subset of the current study population was described in our previous publication. (8) The current analysis included 526 participants with T_2_DM and 1,618 without T_2_DM. While the two groups did not show significant differences in age and sex, the comorbidity profile was significantly higher in the T_2_DM group. The T_2_DM group demonstrated a higher use of AIM (37% vs. 29%, **p**<0.001), and 79% were under antidiabetic medication. Also, significantly higher depression, anhedonia, and melancholia scores were recorded in this group (GDS: 5 vs. 3 points; **p**<0.001); as detailed in another previous publication. (30) Similarly, the T_2_DM group showed a significantly worse cognitive profile (MCI: 19 vs. 17%, Dementia: 9.1% vs. 5.3%; **p**=0.002) and scores (MMSE: 28 vs. 29 points, **p**<0.001; TMT-B: 112 vs. 84 seconds, **p**<0.001). As in the previous publication, T_2_DM participants had significantly higher systemic IL-6 (1.17 vs. 0.88 pg/mL, **p**<0.001) and NfL (16 vs. 13 pg/mL, **p**<0.001) levels.

### 3.2. Correlation between FA in white matter tracts

The correlation between FA values in different WM matter tracts – a surrogate biomarker of structural WM connectivity – was higher in the non-T_2_DM than the T_2_DM group, as demonstrated by the number of significant correlation coefficients across WM FA values in both subgroups (Figure 2).

**Figure 2:**
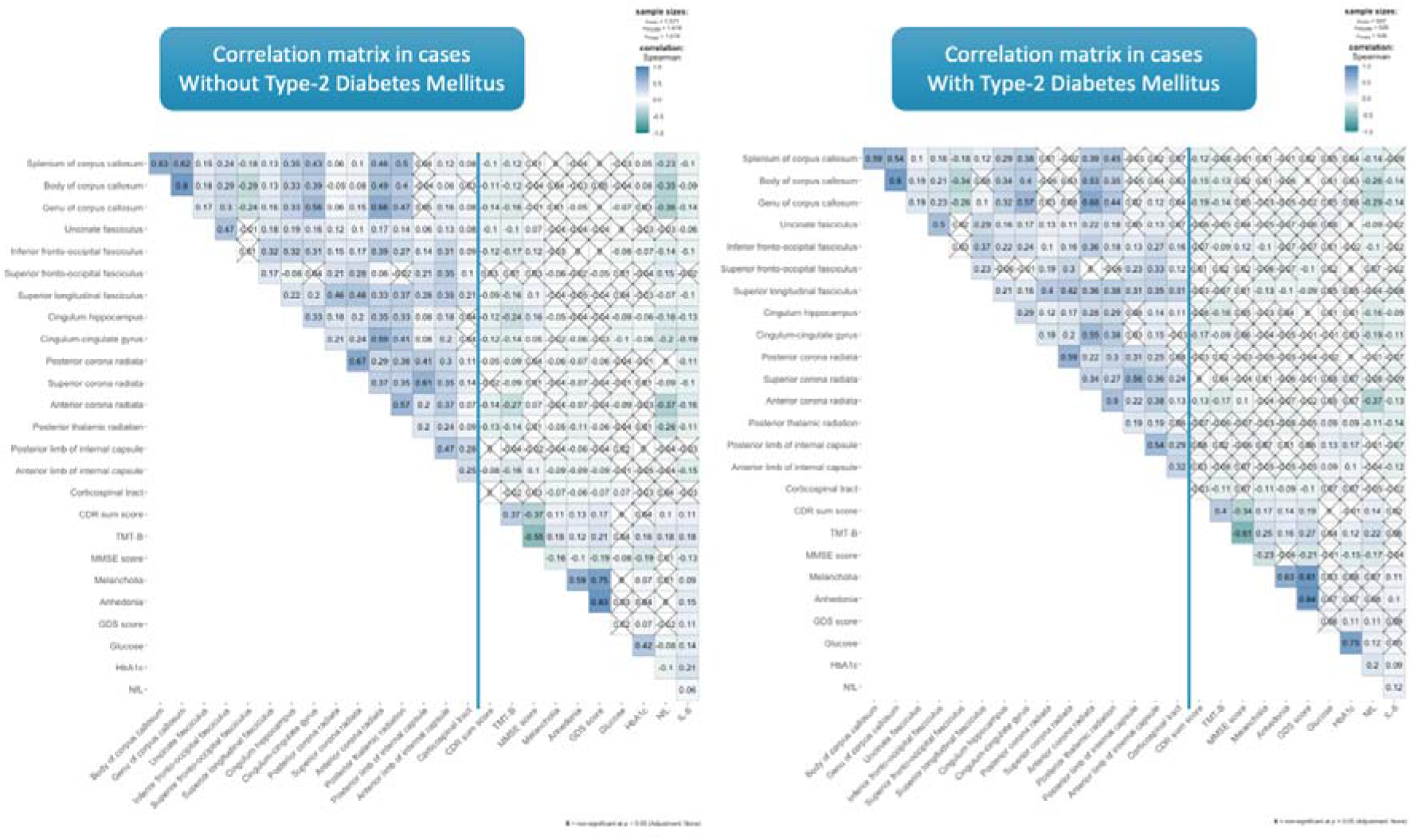
Correlation matrix of immunometabolic biomarkers and fractional anisotropy in different white matter tracts – An averaged approach. 2.a) Cases without type-2 diabetes mellitus. 2.b) Cases with type-2 diabetes mellitus.

### 3.3. Diabetes and white matter integrity

#### 3.3.1. At baseline

After adjusting for the effect of covariates and multiple testing, the non-T_2_DM group showed significantly higher WM FA at baseline, particularly in the Posterior thalamic radiation incl. optic radiation (adj. Cohen’s *d*=0.14 [0.03, 024]), Anterior corona radiata (adj. Cohen’s *d*=0.15 [0.04, 0.26]), Cingulum cingulate gyrus (adj. Cohen’s *d*=0.14 [0.03, 0.25]), Inferior fronto-occipital fasciculus (adj. Cohen’s *d*=0.16 [0.06, 0.27]), Genu of the corpus callosum (adj. Cohen’s *d*=0.13 [0.03, 0.24]), and Body of the corpus callosum (adj. Cohen’s *d*=0.28 [0.17, 0.38]). These will be labeled “T_2_DM-specific WM tracts” for the current study. These effect sizes were small, despite their robust statistical significance. The standardized regression coefficients (*ß*) were comparable with the adjusted effect sizes (adj. Cohen’s *d*), specifically in their statistical significance.

The results remained overall robust after additionally adjusting for the use of AIM (Figure 3).

**Figure 3:**
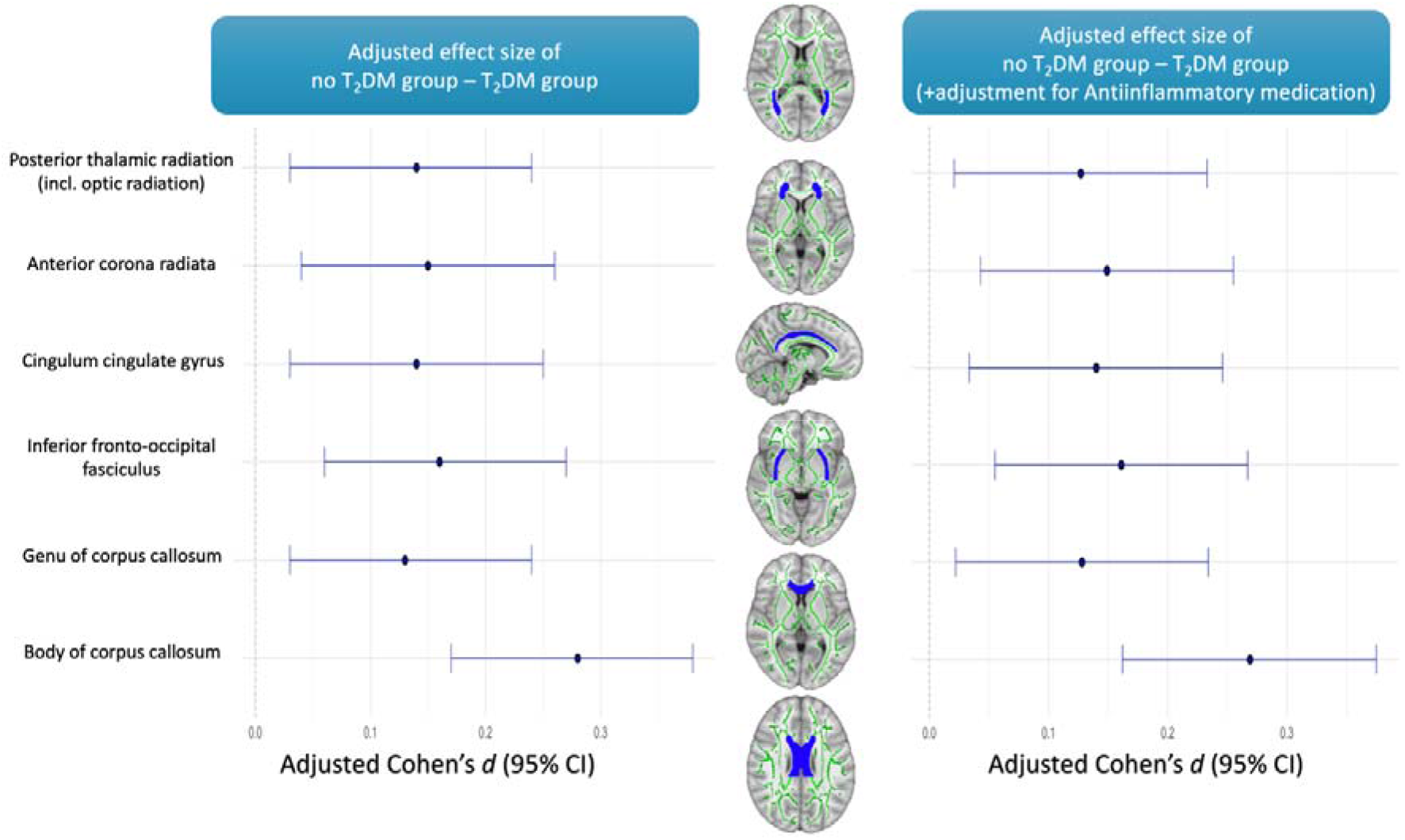
Adjusted effect size of the difference in white matter integrity between participants without and with diabetes: A cross-sectional analysis. 3.a) Without adjustment for the effect of Anti-inflammatory medication. 3.b) After adjustment for the effect of Anti-inflammatory medication.

#### 3.3.2. At two years of follow-up

Despite the lower number at follow-up, the average effect size values observed in the posterior thalamic radiation incl. optic radiation (adj. Cohen’s *d*=0.24 [0.09, 0.39]), anterior corona radiata (adj. Cohen’s *d*=0.23 [0.08, 0.38]), and body of the corpus callosum (adj. Cohen’s *d*=0.30 [0.15, 0.45]) showed an increasing tendency after two years.

No statistically significant difference was observed in the Inferior fronto-occipital fasciculus (adj. Cohen’s *d*=0.11 [-0.04, 0.26]) and genu of the corpus callosum (adj. Cohen’s *d*=0.13 [- 0.02, 0.30]) at follow-up. Differences in the cingulum cingulate gyrus were statistically significant but did not survive the adjustment for multiple testing (adj. Cohen’s *d*=0.17 [0.02, 0.33]).

Differences between the effect sizes observed at baseline and V2 are visualized in Figure 4.

**Figure 4:**
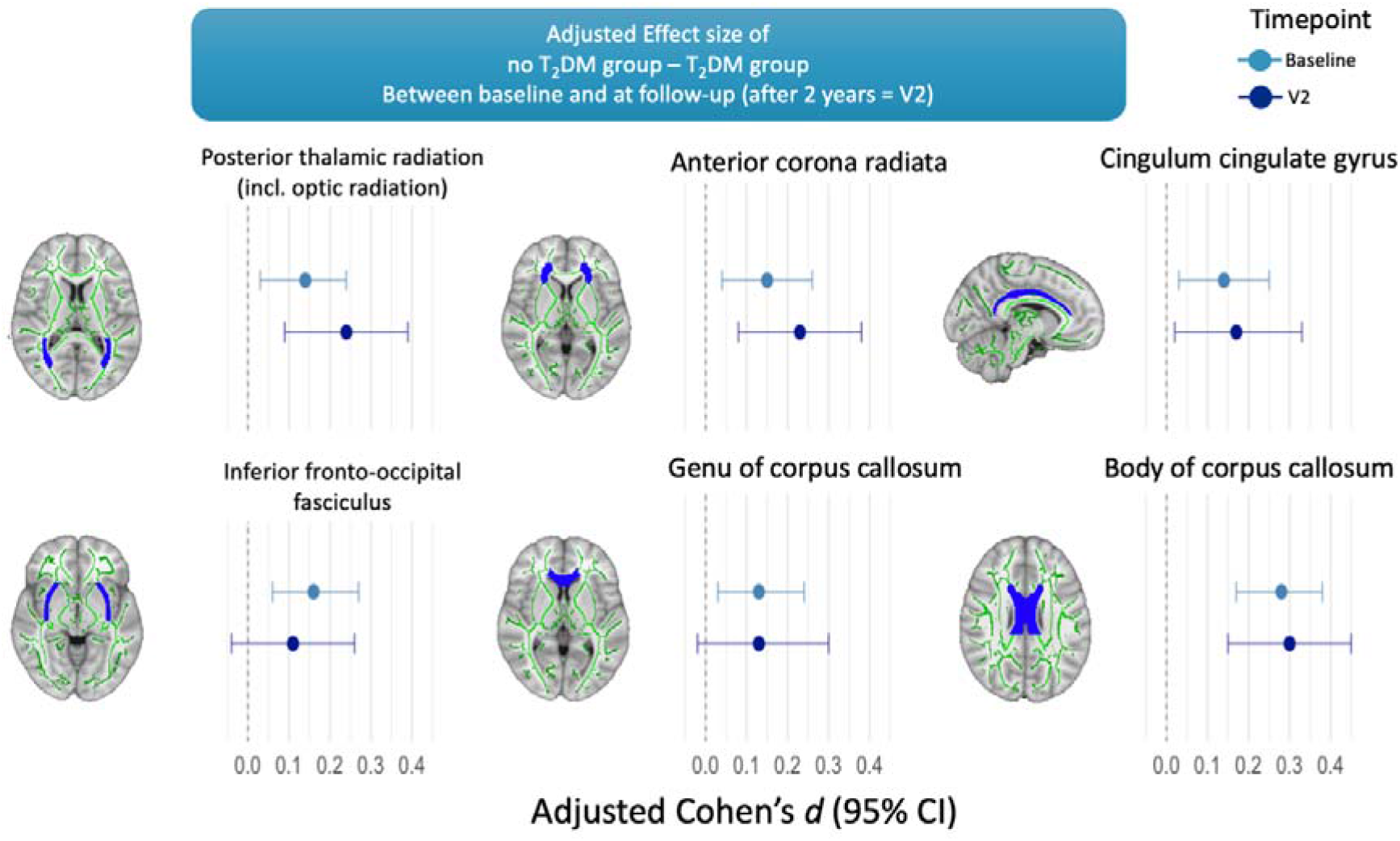
Adjusted effect size of the difference in white matter integrity between participants without and with diabetes: A prospective analysis.

### 3.4. White matter integrity in cognitively unimpaired participants

In the sensitivity analysis including participants without cognitive impairment (HC), WM integrity in the body of the corpus callosum was significantly lower in cases with T_2_DM (adj. Cohen’s *d*=0.25 [0.12, 0.37]), after adjusting for confounders and multiple testing. A similar effect was observed in the inferior fronto-occipital fasciculus (adj. Cohen’s *d*=0.18 [0.06, 0.31]). Details are visualized in Figure 5.

**Figure 5:**
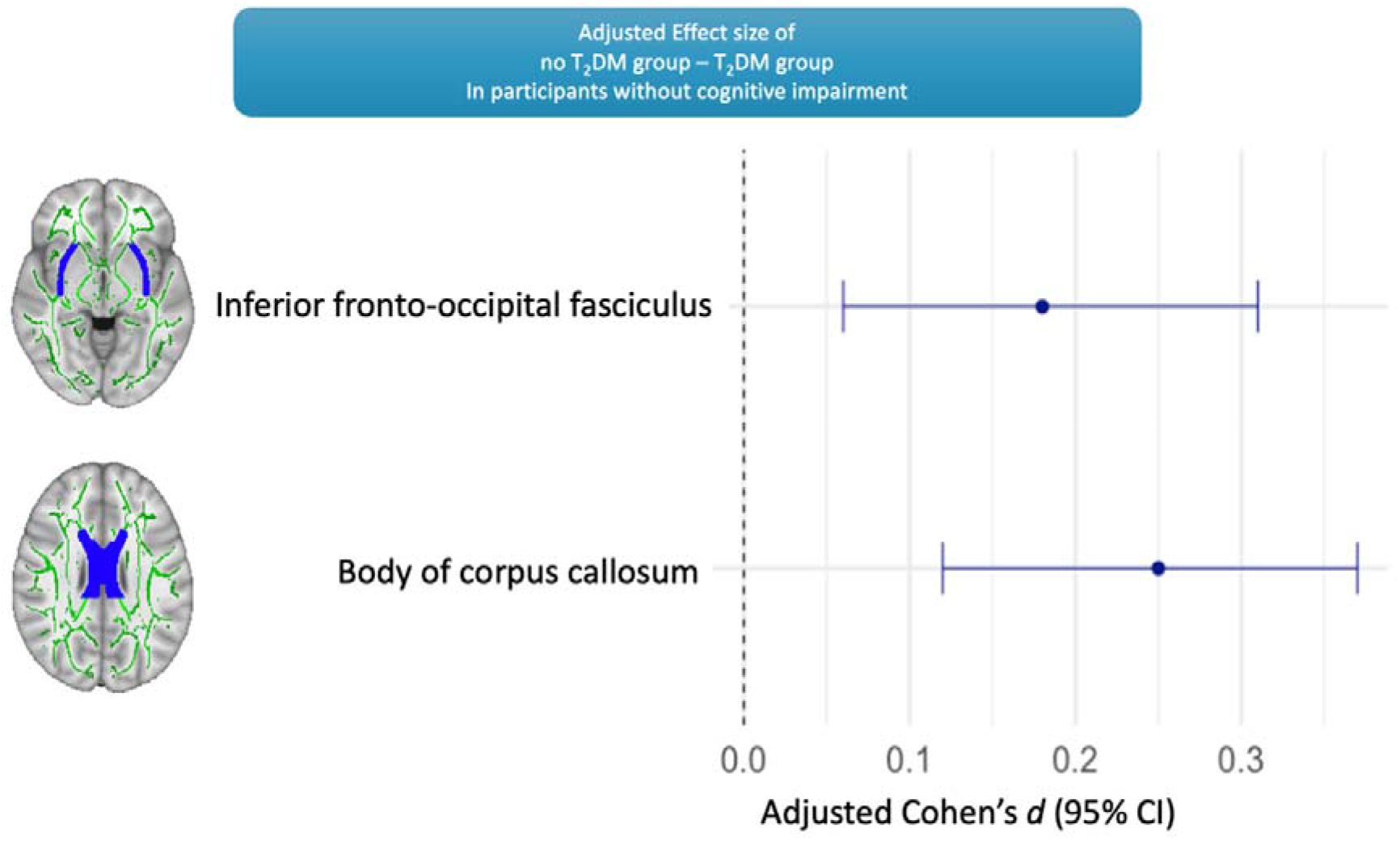
Adjusted effect size of the difference in white matter integrity between cognitively unimpaired participants without and with diabetes.

### 3.5. Mediation analysis

#### 3.5.1. Interleukin-6

Il-6 statistically mediated 13% [4%, 55%] of the association between T_2_DM and FA in the Posterior thalamic radiation incl. optic radiation, 13% [4%, 54%] Anterior corona radiata, 14% [4%, 60%] Cingulum cingulate gyrus, 15% [3%, 81%] Genu of the corpus callosum, and 7% [2%, 14%] Body of the corpus callosum.

No significant mediating effect of IL-6 was observed in the association between T_2_DM and decreased FA in the Inferior fronto-occipital fasciculus. Details are visualized in Figure 6.

**Figure 6:**
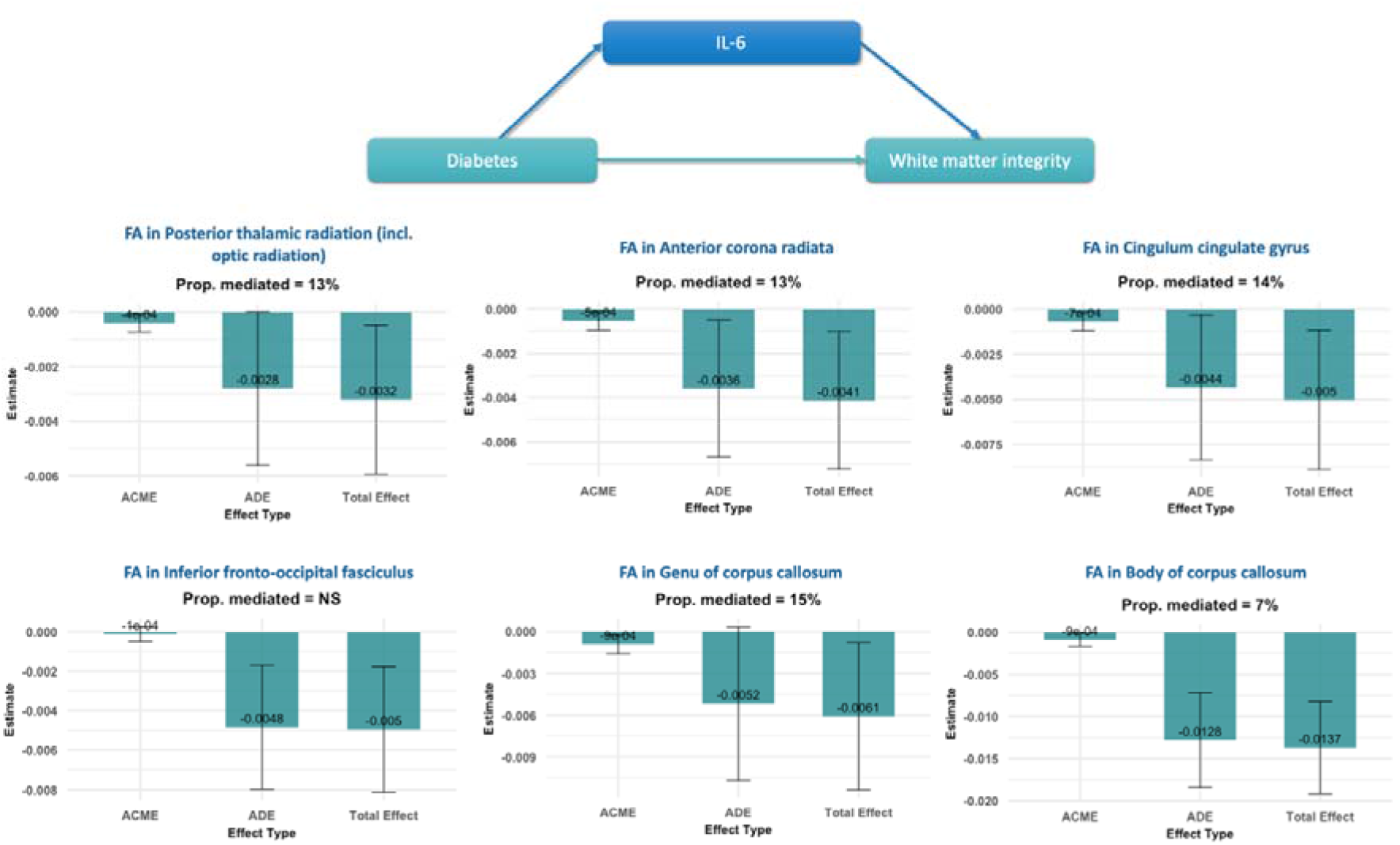
Mediating analysis between diabetes, white matter integrity, and Interleukin-6.

## 4. Discussion

The study showed a significant association between cerebral WM integrity, T_2_DM, and systemic inflammation. Only some WM tracts were specifically affected in T_2_DM participants, despite adjusting for the effect of different biodemographic and comorbid confounders. These WM tracts connect previously described cortical areas affected by neurodegeneration in T_2_DM. The confounder-adjusted effect size in specifically affected WM tracts showed rising patterns within two years of follow-up, despite a prospective decrease in the study population. T_2_DM-specific impairment of WM integrity was objectified in cognitively unimpaired T_2_DM participants. The FA-based correlation matrix showed a decreased in the indirect biomarkers of structural WM connectivity in the T_2_DM group compared to the non-T_2_DM counterpart. The association between WM integrity and T_2_DM was mediated by IL-6, highlighting the role of low-grade inflammation in the T_2_DM-associated intracerebral neuropathy.

### 4.1. White matter integrity and diabetes

#### 4.1.1. WM integrity as an early biomarker

Impairment of WM integrity might start prior to any clinical symptoms and represent, therefore, an early biomarker of neurodegeneration and consequent neurocognitive impairment. (34) In patients with T_2_DM, the WM microstructure of the visual pathways is impaired even in those without diabetic retinopathy. (35) In contrast with previous studies, (34) our study showed that the T_2_DM-specific impairment of WM integrity was not restricted to limbic tracts.

### 4.2. White matter integrity and inflammation

#### 4.2.1. Evidence from clinical studies

Only a few studies explored the association between IL-6 and DTI biomarkers. These included a limited number of patients and showed conflicting results. While some did not find any significant association between IL-6, FA, and mean diffusivity (MD) in a general bi-ethnic population, (36) another study showed significant regional associations between higher IL-6, impaired FA, and radial diffusivity (RD) in schizophrenia patients, but not healthy controls. (37) In a larger sample of participants, there were no genetically explainable correlations between IL-6 levels and DTI parameters on WM microstructures. (33)

Our study showed lower FA in T_2_DM participants compared to the control group. Previous studies in schizophrenia patients showed significant correlations between impaired regional structural integrity of WM, reduced functional connectivity, and higher peripheral inflammation biomarkers (mainly serum IL-6 and CRP). (38)

The association between inflammation and WM integrity is well established in neurological pathologies like multiple sclerosis. (39, 40) Besides, this association has also been described in affective disorders (41) and in severe forms of extra-cerebral physical conditions, such as obstructive sleep apnea and chronic kidney disease. (42, 43) Later studies showed that even in physiological conditions, high or increasing levels of inflammatory biomarkers in early adulthood predict impaired WM structures in later ages. (44) The association between IL-6 levels and WM integrity might start during the conceptional stage. This was shown by the significant association between higher IL-6 levels in women during pregnancy and impaired WM integrity in the uncinate fasciculus of their infants, consequently impacting the cognitive status of the affected ones. (45) Further studies have shown an increased co-occurrence between chronic inflammatory demyelinating neuropathy and diabetes. (46) This supports the hypothesis of shared inflammatory pathways, mainly cytokines, between both pathologies.

The association between inflammation (CRP), poor cognition, and a decreased global FA is particularly significant in APOE ε4 carriers. The affected tracts in a veteran population were the corpus callosum, right superior longitudinal fasciculus, right posterior corona radiata, and the bilateral corona radiata. (47) In a few patients with depression, TNF-alpha and IL-1ß levels were associated with lower FA, particularly in the corpus callosum. (48) Our previous studies highlighted the superiority of IL-6 as a valid biomarker in the immunometabolic brain-body constellation over other inflammatory factors, (30) and this motivated the focus of this analysis on only IL-6. CRP was not measured in African Americans and remained, therefore, out of the scope of the current analysis. However, the production of CRP is regulated by systemic IL-6 levels, and this presents a solid biomarker of CRP variations.

#### 4.2.2. Evidence from preclinical studies

The association between WM integrity and inflammatory biomarkers has been described in preclinical models, where DTI allows the quantification of the effect of inflammation as well as the therapeutic effect of interleukin-1 receptor antagonist (IL-1Ra). (49) The effect of systemic inflammation on cerebral WM integrity starts very early during neurogenesis. Postnatal long-term non-infectious inflammatory states impair the activity of oligodendrocytes and induce WM microstructural anomalies, explaining later developmental deficits in preterm rat infants. (50)

#### 4.2.3. WM integrity, T_2_DM, and alternative diagnostic methods

Diffusion kurtosis imaging (DKI) might provide additional information on T_2_DM-related WM integrity than DTI-specific parameters. Mean kurtosis (MK) values were more sensitive to negative changes associated with T_2_DM disease duration in the genu of the corpus callosum and anterior corona radiata. (51) Although our DTI-based analysis revealed anatomically plausible results, advanced methods such as DKI and neurite orientation dispersion and density imaging (NODDI) might provide additional information.

### 4.3. Clinical implications

Metabolic syndrome - including dyslipidemia and obesity - (52) and modifiable lifestyle habits - including sedentary behavior and smoking - are well-recognized risk factors for peripheral diabetic polyneuropathy (53) and mental health adversities and vice versa. (54) Clinical interventions targeting glucose regulation showed a major role in preventing intracerebral complications. (55) These hypotheses need to be explored in larger randomized controlled trials focusing specifically on WM integrity.

Intensive lifestyle interventions in randomized controlled studies showed beneficial effects in preventing diabetic polyneuropathy. (56) These results might reflect similar preventive effects on intracerebral levels and need to be assessed in longitudinal interventional studies with a special focus on T_2_DM and brain health. This hypothesis is also supported by a Danish population-based study where a decline in diabetic distal symmetrical polyneuropathy incidence in recent years was probably associated with improved medical care. (57) Short-term interventions also showed that physical activity improves the cardiovascular autonomic function in T_2_DM patients, (58) a further potential actor in the interrelation between diabetes and intracerebral WM integrity.

Despite well-monitored T_2_DM, the risk of peripheral diabetic neuropathy tends to increase with the years, (59, 60) and higher triglyceride levels might present a concomitant risk factor that needs to be taken into account in the preventive strategies. (59, 61) However, another study showed that an aggressive lowering of plasma cholesterol levels presents a risk factor for diabetic polyneuropathy. (62) Clinicians need to balance the benefits and risks of T_2_DM-specific and lipid-lowering therapies for better brain health and WM integrity, as well. (63) Initial nerve alterations might be reversible to a certain degree in well-controlled new-onset T_2_DM. (60) For later stages of the disease, a randomized controlled trial showed that intravenous immunoglobulin therapy did not improve the demyelinated peripheral polyneuropathic features in T_2_DM patients. (64) This should motivate early interventional strategies to prevent irreversible complications. Behavioral inflammation-lowering interventions in younger ages (Mediterranean diet, regular physical activity, stress prevention, and mental health support…) might present further strategies to mitigate the T_2_DM-associated inflammatory risks on brain health and, particularly, WM integrity.

### 4.4. Strengths

Peripheral diabetic polyneuropathy is a largely explored pathology, whereas less is known about the involvement of low-grade inflammation in the cerebral WM integrity associated with T_2_DM. This is mainly due to the lack of direct tools to assess WM integrity. dMRI measures are surrogate biomarkers of WM integrity based on water molecule diffusivity. Thus, they provided strong anatomical correlates and are largely used in neuroimaging studies. Second, the cerebral involvement in T_2_DM has so far been less explored, and very little is known about the T_2_DM-brain crosstalk. Such a knowledge gap highlights the novelty of the current study in this evolving area. This is the first study to assess the association between T_2_DM and DTI-related WM integrity with a prospective design, while taking into account the mediating role of IL-6. The high number of participants with equally represented multiethnic backgrounds and with complete biological and neuroimaging data is a major strength of the current study.

### 4.5. Limitations

The study included community-dwelling participants, which might expose the data to selection bias of socially active individuals, and the findings might not be replicated in those with restricted social networks, limited mobility, and poor physical conditions. The study population tends to have an average of a good educational level, and most of the participants with T_2_DM are under at least one form of medication, which does not reflect the findings in less compliant ones and those suffering from severe diabetes complications. Furthermore, the results do not represent potential findings in patients with T_1_DM, who were not eligible for the source cohort. The presented analyses and results do not conclude any causal inference owing to the observational nature of the data. Longitudinal and experimental research is needed to confirm these relationships.

## 5. Conclusions

The study demonstrated T_2_DM-associated impairment of intracerebral WM integrity. As in previous studies, IL-6 seems to play a statistical mediating role in this relationship, partly explaining the observed effects. Its predictive value at early stages of T_2_DM-WM impairment needs to be explored in future studies in order to ensure early preventive interventions. Further longitudinal and experimental studies need to assess causal associations between T_2_DM and WM integrity.

Declarations:

## Ethical approval

All procedures contributing to this work comply with the ethical standards of the relevant national and institutional committees on human experimentation and with the Helsinki Declaration of 1975, as revised in 2013. Ethical approval was obtained from the local institutional review board. Participants gave written informed consent. The current research is based on a secondary analysis of anonymized data and was performed in compliance with the data use agreement (DUA with Hallab).

## Authorship

AH has full access to all of the data and takes responsibility for the integrity of the data and the accuracy of the analysis, visualization, drafting, and editing of the manuscript. Members of the HABS-HD are listed as a group author to acknowledge the collection and provision of the data used in this study, as well as the funding acquisition.

## Data availability

Data can be acquired by qualified researchers after an official request (for more information).

## Data Availability

All data produced in the present study are available upon reasonable request to the authors

